# Biomarkers Correspond with Echocardiographic Phenotypes in Heart Failure with Preserved Ejection Fraction: A Secondary Analysis of the RELAX Trial

**DOI:** 10.1101/2024.04.30.24306660

**Authors:** Peter R. Hyson, David P. Kao

**Affiliations:** University of Vermont Medical Center, Burlington, VT 05401; University of Colorado School of Medicine, Aurora, CO 80045

**Keywords:** heart failure with preserved ejection fraction, latent class analysis, cardiac biomarkers, cystatin-C, NT-proBNP, endothelin-1, high sensitivity troponin, atrial fibrillation

## Abstract

**Background:** Little is known about the relationship between structural phenotypes in in heart failure with preserved ejection fraction (HFpEF) and cardiac biomarkers. We used cluster analysis to identify cardiac structural phenotypes and their relationships to biomarkers in HFpEF.

**Methods and results:** Latent class analysis (LCA) was applied to echocardiographic data including left atrial enlargement (LAE), diastolic dysfunction (DD), E/e’, EF≤55%, and right ventricular dysfunction from 216 patients enrolled in the RELAX trial. Three structural phenotypes were identified. Phenotype A had the most grade II DD. Phenotype B had the most grade III DD, worst LAE, elevated E/e’ and right ventricular dysfunction. Phenotype C had the least DD and moderate LAE. Phenotypes B and C had prevalent atrial fibrillation (AF). Phenotype B patients had increased carboxy-terminal telopeptide of collagen type I (CITP), cystatin-c (CYSTC), endothelin-1 (ET1), NT-proBNP, and high-sensitivity troponin I (TNI). Type A had the next highest CITP and CYSTC levels while Type C had next highest NT-proBNP.

**Conclusions:** Structural HFpEF phenotypes demonstrated different characteristics including cardiac biomarkers. These findings may help explain phenotype-specific differences in natural history and prognosis, and they may represent phenotype-specific pathophysiology that could be amenable to targeted therapy.

## INTRODUCTION

Heart failure with preserved ejection fraction (HFpEF) accounts for nearly half of all heart failure cases (1). Despite several large randomized controlled trials, no medical treatment for HFpEF has demonstrated a convincing therapeutic benefit. HFpEF is generally defined as a clinical syndrome which presents with the signs and symptoms of heart failure but with ejection fraction (EF) >50%. Rather than comprising a single pathophysiology and natural history of disease, HFpEF is a heterogeneous disorder with differing phenotypic characteristics, pathophysiology and prognosis. This suggests that targeting specific therapies to individual HFpEF phenotypes might offer success where one-size-fits-all approaches have failed (2).

One promising approach to subdividing HFpEF into targetable subgroups is through use of unsupervised machine learning techniques such as cluster-based analysis. Some analyses have combined continuous echocardiographic measurements with other elements (e.g. blood count components, electrolytes, electrocardiogram findings) (3, 4). However thus far cluster-based methods approach have not examined the association of structural HFpEF phenotypes with biomarkers or exercise capacity.

The Effect of Phosphodiesterase-5 Inhibition on Exercise Capacity and Clinical Status in Heart Failure with Preserved Ejection Fraction (RELAX) trial studied the impact of sildenafil vs. placebo on peak oxygen requirement in patients with HFpEF (5). The investigators collected echocardiographic findings, maximum oxygen consumption (VO2), 6-minute walk performance, and biomarker levels including endothelin 1(ET1), NT-proBNP, carboxy-terminal telopeptide of collagen type 1 (CITP), cystatin-c (CYSTC), and high-sensitivity troponin I (TNI). These biomarkers have been previously found to be relevant to HFpEF with ET1, TNI, and NT-proBNP prognosticating poor outcomes (6–8), CITP predictive of incident HFpEF (9), and CYSTC associated with diastolic dysfunction (10). However the association of these biomarkers relative to complex HFpEF structural phenotypes is unknown.

Our goal was to apply cluster-based analysis to imaging data from RELAX to elucidate complex cardiac structural phenotypes. We hypothesized that cardiac structural phenotypes are associated with distinct biomarker profiles, which might provide insight into underlying pathophysiology.

## METHODS

The design and findings of the RELAX trial have been described previously (5). Briefly, RELAX was a multicenter, double-blind, randomized, placebo-controlled trial of 216 outpatients with HFpEF (EF > 50%), reduced exercise capacity (60% or less of age/sex-specific normal value with a respiratory exchange ratio of 1.0 or more) and elevated NT-proBNP or elevated filling pressures. Mean total follow-up time was 5.3 months. The primary outcome was change in peak oxygen requirement (VO2) at 24 weeks with secondary endpoints including change in 6-minute walk distance and change in a composite clinical status score based on outcomes and symptom burden. Sildenafil was not associated with improvement in any of the primary or secondary outcomes.

Structural phenotype definitions were derived using latent class analysis (LCA) to identify patterns of five echocardiographic features:□left atrial enlargement (LAE),□diastolic dysfunction (DD),□medial E/e’, ejection fraction (EF) greater or less than 55%, and right ventricular□function (Table 1). Left ventricular mass index was not included in LCA, as 92% fell within the normal range adjusted for sex (11). The optimal number of phenotypes was determined using the Akaike Information Criterion and adjusted Bayesian Information Criterion. Clinical characteristics including biomarkers were compared between phenotypes using chi-square (categorical) and Kruskal-Wallis (continuous) tests. Outcomes included change in peak VO2 max, six-minute walk, Minnesota Living With Heart Failure (MWLHF) total score, and changes in serum aldosterone, high sensitivity-C reactive protein (hs-CRP), CITP, CYSTC, ET1, Galactin-3, NT-proBNP, PIIINP, and hs-TNI. Outcomes were compared between treatment arms within each phenotype. A p-value < 0.05 was considered significant throughout. All statistical analysis was performed using the R statistical package (version 3.6.3, R Foundation for Statistical Computing, Vienna, Austria) and RStudio (version 1.2.5033, RStudio, Inc., Boston, MA). LCA was performed using the *poLCA* library (12).

**Table 1a.** Latent class analysis component echo parameters by structural phenotype class.

|  | Type |  |  | Combined | P-value |
| --- | --- | --- | --- | --- | --- |
|  | A | B | C |  |  |
|  | (N=45) | (N=103) | (N=68) | (N=216) |  |
| LA volume index, ml/m<sup>2</sup> |  |  |  |  |  |
| < 34 | 10 (31) | 6 (9) | 13 (28) | 29 (19) | 0.002 |
| 34-41 | 10 (31) | 8 (11) | 8 (17) | 26 (17) |  |
| 41-48 | 5 (16) | 14 (20) | 8 (17) | 27 (18) |  |
| > 48 | 7 (22) | 42 (60) | 18 (38) | 67 (45) |  |
| Diastolic dysfunction |  |  |  |  |  |
| Normal | 0 (0.0) | 25 (32) | 55 (86) | 80 (43) | <0.001 |
| Grade I | 6 (14) | 0 (0.0) | 5 (8) | 11 (6) |  |
| Grade II | 36 (86) | 0 (0.0) | 2 (3) | 38 (21) |  |
| Grade III | 0 (0) | 54 (68) | 2 (3) | 56 (30) |  |
| Septal E/e' > 15 | 32 (74) | 75 (91) | 0 (0) | 107 (57) | <0.001 |
| RV function dysfunction |  |  |  |  |  |
| Normal | 42 (98) | 71 (72) | 54 (83) | 167 (81) | 0.003 |
| Mild | 0 (0.0) | 14 (14) | 8 (12) | 22 (11) |  |
| Moderate | 0 (0.0) | 13 (13) | 3 (5) | 16 (8) |  |
| Severe | 1 (2) | 0 (0) | 0 (0) | 1 (0) |  |
| LV ejection fraction, % | 65 [60,68] | 58 [55,65] | 60 [55,65] | 60 [55,65] | <0.001 |
| LA = Left atrial LV = Left ventricular RV = Right ventricular |  |  |  |  |  |

**Table 1b.**
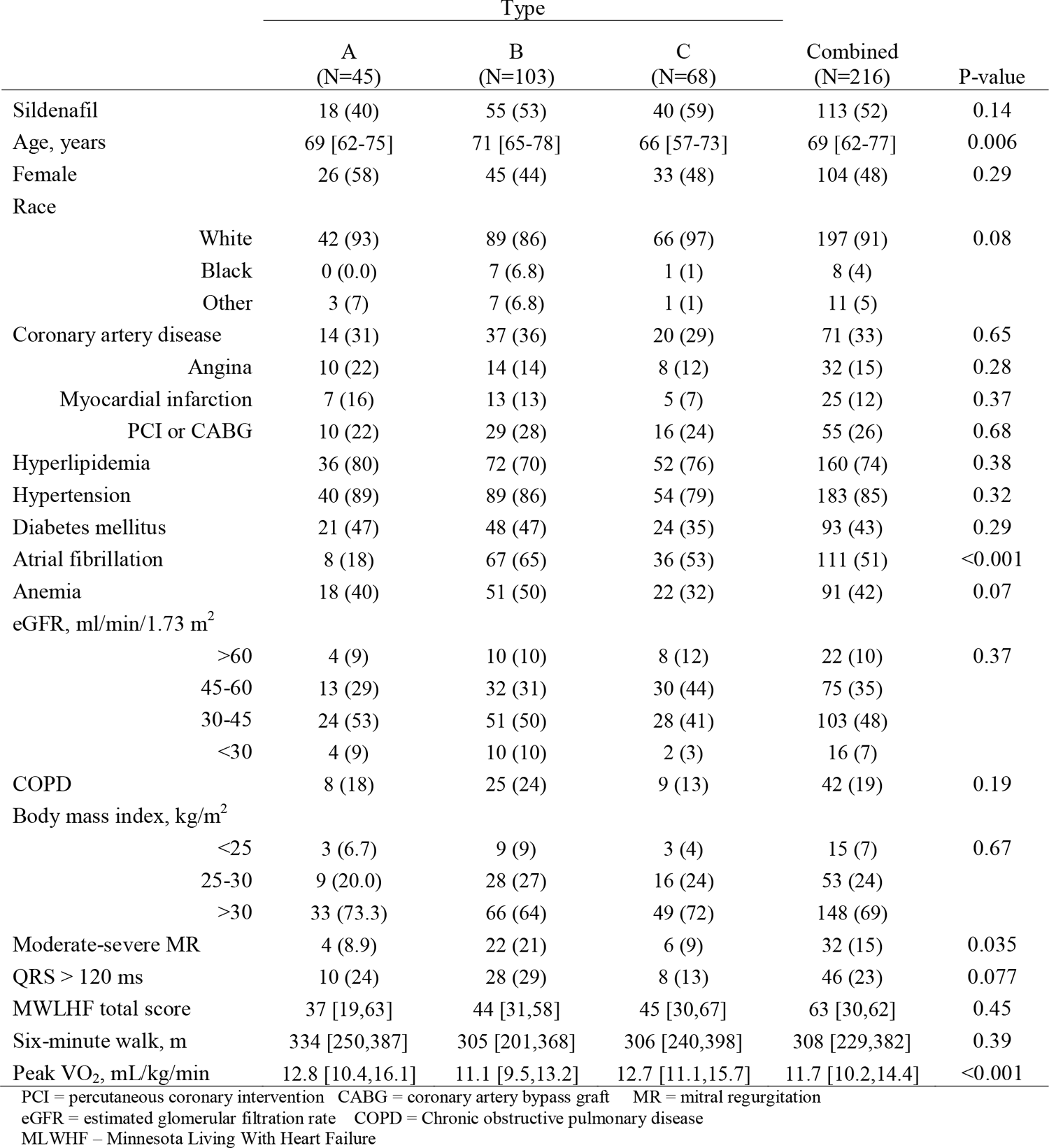
Clinical characteristics of structural phenotypes,, N (%) or median [interquartile range].

## RESULTS

We identified three structural phenotypes, which differed significantly across all 5 LCA variables (**Table 1a**). Phenotype A was characterized by moderate (grade I—II) DD, the lowest rate of LAE, the least right ventricular dysfunction, and the highest EF. Phenotype B, the largest of the three subgroups, had the highest rate of grade III DD, the most LAE, the most elevated E/e’, and the most right ventricular dysfunction. Phenotype C subjects had little to no DD but had the second highest rate of LAE, and normal E/e’. Although not included in LCA, right ventricular systolic pressure was also significantly higher in Phenotype B (median 48 mm Hg [interquartile range (IQR) 38-55]) compared with Phenotype A (median 36 mm Hg [IQR 32-43]) and Phenotype C (median 36 mm Hg [IQR 29-47]). Clinical characteristics associated with each structural phenotype are also summarized in **Table 1b**.

Phenotype B was significantly older (median 71 years [interquartile range (IQR) 65-78]) compared with Phenotype A [median 69 years [IQR 62-75]) and Phenotype C (median 66 years [IQR 57-73]). The three groups did not have significantly different distributions of race or sex and no significant difference in prevalence of coronary artery disease, hypertension, diabetes, COPD, or CKD. Significant differences between groups were observed in LVEF, where Phenotype B had the lowest and Phenotype A highest, and atrial fibrillation (AF) where Phenotypes B and C both had a high prevalence at 65% and 53% respectively compared with 18% in Phenotype A (p<0.001). There were no significant differences between phenotypes in total Minnesota Living With Heart Failure score or 6 minute walk test, but Phenotype B subjects had significantly lower VO2 max at baseline (median 11.1 ml/kg/min [IQR 9.5-13.2]) than Phenotype A (median 12.8 ml/kg/min [IQR 10.4-16.1]) and C (median 12.7 ml/kg/min [IQR 11.1-15.7], p<0.001).

CITP, CYSTC, ET1, NT-BNP, and TNI were significantly different at baseline across the three phenotypes (**Table 2**). Phenotype B had the highest baseline levels of CITP, CYSTC, ET1, NT-proBNP, and TNI, whereas subjects in Phenotype C had the lowest levels of CITP, CYSTC, ET1, and TNI. Baseline cyclic GMP did not differ significantly between groups.

**Table 2.** Baseline biomarker levels by echo structural phenotype class, median [interquartile range].

|  | Type A<br>(N=45) | Type B<br>(N=103) | Type C<br>(N=68) | Combined<br>(N=216) | P-value |
| --- | --- | --- | --- | --- | --- |
| Aldosterone, ng/dL | 206 [112,263] | 207 [123,285] | 168 [119,297] | 189 [119,282] | 0.69 |
| hs-CRP, mg/L | 3.8 [2.0,7.2] | 3.6 [1.9,8.3] | 3.8 [1.6,8.3] | 3.7 [1.8,8.1] | 0.84 |
| cGMP, pmol/mL | 75 [55,112] | 76 [58,95] | 82 [61,106] | 77 [57,101] | 0.53 |
| CITP, µg/L | 6.0 [4.4,9.8] | 6.7 [5.4,10.6] | 5.7 [4.1,8.2] | 6.3 [4.7,9.9] | 0.031 |
| Cystatin C, mg/L | 1.33 [1.06,1.76] | 1.45 [1.14,1.83] | 1.15 [0.93,1.53] | 1.30 [1.07,1.74] | 0.002 |
| Endothelin-1, pg/mL | 2.20 [1.82,2.43] | 2.85 [2.16,3.70] | 2.14 [1.84,2.71] | 2.36 [1.95,3.20] | <0.001 |
| Galectin-3, ng/mL | 13.6 [11.7,16.8] | 14.2 [11.1,18.6] | 13.6 [11.2,17.0] | 13.8 [11.1,17.8] | 0.8 |
| NT-proBNP, ng/L | 441 [184,693] | 1171 [531,2032] | 481 [91,1237] | 700 [283,1553] | <0.001 |
| PIIINP, µg/L | 6.9 [5.3,8.9] | 7.9 [6.3,10.7] | 7.7 [6.0,9.2] | 7.7 [6.1,10.0] | 0.2 |
| hs-Troponin I, pg/mL | 8.3 [4.9,15.3] | 11.6 [6.9,35.4] | 6.7 [3.6,12.4] | 9.5 [5.4,19.7] | <0.001 |
| hs-CRP = high sensitivity C-reactive protein cGMP = cyclic guanosine monophosphate<br>CITP = carboxy-terminal telopeptide of type I collagen NT-proBNP = N-terminal proB-type natriuretic peptide PIIINP =<br>procollagen III amino terminal propeptide |  |  |  |  |  |

Outcomes according to structural phenotype are summarized in **Table 3**. No significant changes were observed in MLWHF score or exercise capacity between treatment arms in any phenotype. In Phenotype B sildenafil-treated subjects had significantly greater increases in levels of ET1 (0.4 vs.-0.0005, p=0.004) and uric acid (0.4 vs. -0.3, p=0.03) at 24 weeks compared with placebo. Otherwise, there were no significant differences in biomarker level changes between treatment arms within the three structural phenotypes.

**Table 3.**
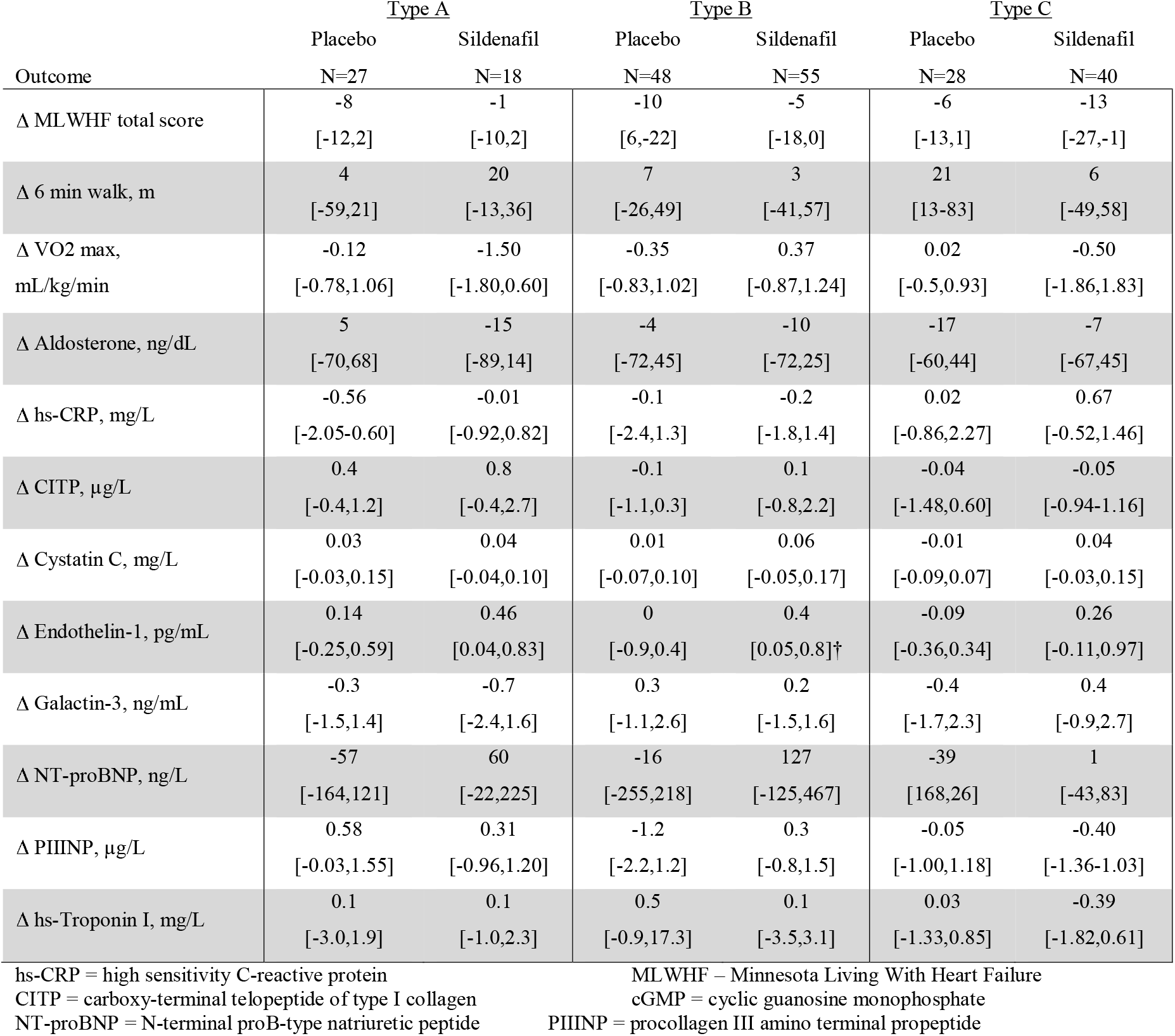
Outcomes by structural phenotype, median [interquartile range].

| Outcome | Type A |  | Type B |  | Type C |  |
| --- | --- | --- | --- | --- | --- | --- |
|  | Placebo<br>N=27 | Sildenafil<br>N=18 | Placebo<br>N=48 | Sildenafil<br>N=55 | Placebo<br>N=28 | Sildenafil<br>N=40 |
| Δ MLWHF total score | -8<br>[-12,2] | -1<br>[-10,2] | -10<br>[6,-22] | -5<br>[-18,0] | -6<br>[-13,1] | -13<br>[-27,-1] |
| Δ 6 min walk, m | 4<br>[-59,21] | 20<br>[-13,36] | 7<br>[-26,49] | 3<br>[-41,57] | 21<br>[13-83] | 6<br>[-49,58] |
| Δ VO2 max,<br>mL/kg/min | -0.12<br>[-0.78,1.06] | -1.50<br>[-1.80,0.60] | -0.35<br>[-0.83,1.02] | 0.37<br>[-0.87,1.24] | 0.02<br>[-0.5,0.93] | -0.50<br>[-1.86,1.83] |
| Δ Aldosterone, ng/dL | 5<br>[-70,68] | -15<br>[-89,14] | -4<br>[-72,45] | -10<br>[-72,25] | -17<br>[-60,44] | -7<br>[-67,45] |
| Δ hs-CRP, mg/L | -0.56<br>[-2.05-0.60] | -0.01<br>[-0.92,0.82] | -0.1<br>[-2.4,1.3] | -0.2<br>[-1.8,1.4] | 0.02<br>[-0.86,2.27] | 0.67<br>[-0.52,1.46] |
| Δ CITP, μg/L | 0.4<br>[-0.4,1.2] | 0.8<br>[-0.4,2.7] | -0.1<br>[-1.1,0.3] | 0.1<br>[-0.8,2.2] | -0.04<br>[-1.48,0.60] | -0.05<br>[-0.94-1.16] |
| Δ Cystatin C, mg/L | 0.03<br>[-0.03,0.15] | 0.04<br>[-0.04,0.10] | 0.01<br>[-0.07,0.10] | 0.06<br>[-0.05,0.17] | -0.01<br>[-0.09,0.07] | 0.04<br>[-0.03,0.15] |
| Δ Endothelin-1, pg/mL | 0.14<br>[-0.25,0.59] | 0.46<br>[0.04,0.83] | 0<br>[-0.9,0.4] | 0.4<br>[0.05,0.8]† | -0.09<br>[-0.36,0.34] | 0.26<br>[-0.11,0.97] |
| Δ Galactin-3, ng/mL | -0.3<br>[-1.5,1.4] | -0.7<br>[-2.4,1.6] | 0.3<br>[-1.1,2.6] | 0.2<br>[-1.5,1.6] | -0.4<br>[-1.7,2.3] | 0.4<br>[-0.9,2.7] |
| Δ NT-proBNP, ng/L | -57<br>[-164,121] | 60<br>[-22,225] | -16<br>[-255,218] | 127<br>[-125,467] | -39<br>[168,26] | 1<br>[-43,83] |
| Δ PIIINP, μg/L | 0.58<br>[-0.03,1.55] | 0.31<br>[-0.96,1.20] | -1.2<br>[-2.2,1.2] | 0.3<br>[-0.8,1.5] | -0.05<br>[-1.00,1.18] | -0.40<br>[-1.36-1.03] |
| Δ hs-Troponin I, mg/L | 0.1<br>[-3.0,1.9] | 0.1<br>[-1.0,2.3] | 0.5<br>[-0.9,17.3] | 0.1<br>[-3.5,3.1] | 0.03<br>[-1.33,0.85] | -0.39<br>[-1.82,0.61] |

hs-CRP = high sensitivity C-reactive protein
CITP = carboxy-terminal telopeptide of type I collagen
NT-proBNP = N-terminal proB-type natriuretic peptide
MLWHF – Minnesota Living With Heart Failure
cGMP = cyclic guanosine monophosphate
PIIINP = procollagen III amino terminal propeptide

## DISCUSSION

Application of cluster analysis to echocardiographic data from HFpEF patients reveals structural phenotypes which are characterized by diastolic dysfunction of varying severity, in the case of Phenotypes A and B, and atrial fibrillation and LAE without other major abnormalities in the case of Phenotype C. Structural and clinical characteristics Phenotype B are congruent with classic descriptions of HFpEF as a disease of the elderly characterized by severe DD complicated by AF and myocardial fibrosis. Phenotype B clearly represents the most severe structural HFpEF phenotype in this dataset with Phenotype C standing in dramatic contrast in terms of both echocardiographic findings and biomarker profile. Phenotype B, which had the highest median age and highest prevalence of AF also had the highest levels of CITP, CYSTC, ET1, NTBNP, and TNI. Phenotype C, with the youngest patients, had significantly lower levels of these biomarkers. This constellation of findings lends credence to the concept of different structural phenotypes having differences in underlying pathophysiology. Furthermore, although Phenotype B subjects were older than those in the other two groups, other baseline characteristics were not significantly different between groups. We therefore believe that differences in biomarkers are primarily linked to echocardiographic findings rather than other clinical features. Sildenafil had little to no effect irrespective of cardiac structural phenotype, suggesting that pathophysiologic differences between phenotypes did not impact response to phosphodiesterase-5 inhibition.

Factors apart from cardiac structure such as AF or non-cardiac contributors can produce symptoms consistent with HF. Phenotype C had comparatively few structural abnormalities and the lowest E/e’ of the 3 phenotypes, calling into question the cause of HF symptoms in this population. The most notable abnormalities were the relatively high rate of severe LAE (38%) and AF (53%). NT-proBNP was elevated in Phenotype C in the setting of prevalent LAE, but not significantly different than Phenotype A (p=0.91). However the NT-proBNP level in Phenotype C was markedly lower than Phenotype B (median 481 [IQR 91-1237] vs. 1171 [IQR 531-2032], p<0.001). While LAE is an important finding in DD, the fact that this population did not have echocardiographic evidence of DD and did not show elevated CYSTC argues for a different pathophysiologic mechanism for clinical HFpEF. Given the 100% normal E/E’ and comparatively low RVSP, it is possible that this structural phenotype reflects younger patients with HF symptoms and functional limitations driven by primary rather than secondary AF. In such patients, medical therapies directed at ventricular remodeling or attenuating diastolic dysfunction could be less effective. Instead, therapies directed at heart rhythm control such as antiarrhythmic therapy or catheter ablation may prove more beneficial (13).

Phenotype B patients had cardiac structural abnormalities accompanied by elevation of several biomarker elevations including CYSTC, NT-proBNP, TNI, CITP, ET1. The mechanism of the relationship between these biomarker abnormalities and structural phenotype B is unclear. The biomarker abnormalities may reflect an underlying molecular mechanism leading to structural change, or conversely, primary structural abnormalities lead to changes in serum biomarkers. One common denominator among several of the above biomarkers of interest is their role in cardiac remodeling and diastolic dysfunction. Phenotype A typified by moderate DD showed elevations in CYSTC, CITP, ET1, TNI compared with Phenotype C, although these were smaller in magnitude compared with Phenotype B. We believe that this phenotype reflects some shared structural pathophysiology to Phenotype B although the marked difference in rate of AF (18% vs. 64%) suggests that Phenotype A may not simply be a less severe or early stage of Phenotype B.

CYSTC is an inhibitor of cysteine proteases called cathepsins which degrade the extracellular matrix, which has been found to be a marker of diastolic dysfunction and to be associated with increasing LAE (10, 14). These associations may be mediated through altered collagen metabolism resulting in myocardial fibrosis (15). Cathepsins and CYSTC may be elevated in rats and humans with hypertension-induced left ventricular hypertrophy (16, 17). CITP and ET1 also appear to be associated with cardiac remodeling. CITP is a collagen metabolite and like CYSTC, increased CITP has been observed with cardiac fibrosis and diastolic dysfunction (18). Likewise, ET1 is part of a signaling pathway in which myocyte stress or injury leads to myocyte hypertrophy (19).

One possible etiology for disease in Phenotypes A and B is microvascular dysfunction. PROMIS-HFpEF showed that even in a cohort of HFpEF patients that excluded unrevascularized coronary artery disease, prevalence of microvascular coronary dysfunction as measured by coronary flow reserve approaches 75% (20). This may contribute to a lack of functional reserve and metabolic demand noted above. In addition to collagen metabolism and cardiac remodeling, CYSTC is also associated with cardiac injury. In rat cardiac myocytes, release of CYSTC may be provoked by exposure to reactive oxygen species, implying a role for myocyte injury leading to cardiac remodeling (22). This syndrome of microvascular dysfunction, lack of ventricular reserve, diastolic dysfunction due to remodeling and myocardial stiffening may have its roots in metabolic causes: study in swine has shown that similar findings can be provoked by induced hypertension, diabetes, and hyperlipidemia (23).

Phenotype B’s relative elevations in TNI and ET1 are also indicators of disease severity. TNI has been shown to prognosticate all-cause mortality and rehospitalization in stable HFpEF whereas ET1 has been found to be both predictive of future hospitalizations and indicative of incipient pulmonary hypertension (6, 7). TNI elevation may reflect ongoing myocyte injury secondary to interplay between diastolic dysfunction and poor ventricular functional reserve in which increasing oxygen demand is unmatched by supply (24). LAE might also give the type B patient a greater likelihood of developing secondary AF which was also quite prevalent in this group (65%).

NT-proBNP, is associated with increasing left atrial enlargement and E/e’ (and implicitly HFpEF) but is not known to play a direct role in myocardial fibrosis (25). Rather, NT-proBNP is synthesized in response to both atrial and ventricular wall stress and appears to be protective against remodeling (26, 27). NT-proBNP elevation in Phenotype B could be interpreted more as myocardial response to stress (be it from increased filling pressures, afterload, etc) then as a component of a remodeling pathway.

Phenotype B, with its evident severity of echocardiographic findings as well as biomarker elevation appears to be the best candidate for novel medical therapies directed at ventricular remodeling. Endothelin receptor antagonism and neprilysin inhibition are two therapies that may be particularly effective in this subset of HFpEF.Although initial trial of endothelin receptor antagonism in HFpEF generally has thus far been unsuccessful, murine models have been encouraging, and perhaps targeted recruitment of a cohort of patients resembling those in Phenotype B would yield a more promising result than in prior investigations (28, 29). Similarly, PARAGON-HF showed some evidence of benefit for sacubitril-valsartan in subgroups with lower EF and in women-perhaps application of LCA to this study group might reveal a therapeutic signal within a cohort resembling Phenotype B (30).

### Limitations

This was a retrospective, secondary analysis and thus all findings must be validated prospectively. Having subdivided the sample into three, this analysis was likely underpowered to detect a significant difference associated with treatment between subsets. LCA is well-suited to categorization of multiple interacting clinical observations (e.g. DD, sex-specific definitions of normal/abnormal), it does limit ability to assess magnitude of association with continuous variables with different phenotypes, and data-driven methods of selecting the ideal component variables apart from expert opinion remain in evolution.

### Conclusion

Complex HFpEF structural phenotypes exist which are associated with distinct patterns of biomarker expression. The combination of structural phenotype, biomarker abnormalities, and clinical profiles may help identify effective therapies targeted to specific HFpEF phenotypes.

## Data Availability

All primary data used in the present study are available from the NHLBI BioLINCC resource by application. Harmonized data derived from BioLINCC data are available from the authors on reasonable request.

https://biolincc.nhlbi.nih.gov/studies/hfn_relax/

